# WakeSentry: real-time awakening detection from single-channel EEG across sleep cohorts and forehead headbands

**DOI:** 10.64898/2026.11.17.26368304

**Authors:** Han Cui, Yuge Zhu, Bing Li, Shaojia Huang, Tao Xu

**Author notes:** Corresponding authors: Bing Li, Department of Electronic Engineering, College of Engineering, Shantou University, 243 Daxue Road, Shantou 515063, China; Tao Xu, Department of Biomedical Engineering, College of Engineering, Shantou University, 243 Daxue Road, Shantou 515063, China; Shaojia Huang, School of Intelligent Manufacturing and Aeronautics, Zhuhai College of Science and Technology, 8 Anji East Road, Jinwan District, Zhuhai 519041, China.

## Abstract

**Objective/Background:** Awakenings after sleep onset are a core measure of sleep continuity, and EEG headbands and closed-loop stimulators, now used at home, must detect each one as it happens, to count it or to stop stimulation. Open sleep stagers match expert scorers, yet have been evaluated only offline over whole nights, so their real-time performance is untested.

**Patients/Methods:** To fill this gap, we developed WakeSentry, which reads only past frontal EEG and learns the moment of waking through an awakening module. We tested it on 1,529 nights from five polysomnography (PSG) cohorts, three clinical, each held out of training, and on 224 headband nights, against GSSC, U-Sleep and YASA.

**Results:** At a matched rate of 0.5 false awakenings per hour, WakeSentry caught more awak-enings than GSSC in every cohort and on both headbands. On PSG its sensitivity rose from 63.8% to 70.5%, above U-Sleep and YASA on average. On the headbands it rose from 49.8% to 53.9%, a gain lost without the awakening module. With thresholds preset on the training cohorts, it still outperformed GSSC in six of seven test sets, with at most 0.40 false awakenings per hour. As a replayed stimulator gate, it shortened stimulation during wake in five test sets while staying open for at least 85% of N2 and N3.

**Conclusions:** WakeSentry therefore works in patients and on unseen headbands. It can count awakenings at home and stop stimulation at each one, enabling multi-night monitoring of insomnia and closed-loop stimulation that stops when the sleeper wakes.

## 1. Introduction

Waking during the night fragments sleep, and fragmented sleep has a cost the next day. Although healthy sleepers roused by brief sounds every 2 min slept as long as on undisturbed nights, they were sleepier the next day and did worse on tests of attention and mental flexibility [1]. Clinically, this fragmentation is tracked as wake after sleep onset, a core measure of sleep continuity that, like brief arousals, increases with age [2,3]. Awakenings also lie at the centre of insomnia, which is still diagnosed from clinical interviews and sleep diaries [4]. In insomnia, wake after sleep onset varies widely from one night to the next [5], so a single laboratory night describes it poorly and repeated measurement at home is needed.

Devices that work through the night can now make this measurement, provided they recognise awakenings as they occur. Wrist actigraphy and consumer trackers detect sleep well but wake poorly [6,7]; against polysomnography, actigraphy labelled only about one third of wake epochs as wake [6]. EEG recorded on the forehead or around the ear follows sleep stages more closely. The automatic staging of a dry-EEG headband agreed with expert scoring of PSG on 83.5% of epochs, close to the 86.4% that individual experts reached [8], and single-channel forehead and around-the-ear EEG have also been compared with PSG in clinical and research cohorts [9–11]. Such headbands now record sleep at home over many nights [12,13], and to report sleep continuity they must count awakenings. Closed-loop stimulation asks more of them, because it acts on the sleeping brain. Sounds timed to slow oscillations enhance slow-wave activity and memory consolidation in young [14] and older adults [15], and the method has moved to headbands used at home on consecutive nights [16–18]. Memory cues replayed in NREM sleep strengthen the cued memories, whereas the same cues given during wake do not [19,20]. A stimulator has to stop as soon as the sleeper wakes, since sounds that reach a waking sleeper add to the fragmentation that stimulation should avoid [1], and stopping on time requires scoring sleep as it happens [21].

Automatic sleep staging now reaches the agreement of human scorers on PSG [22,23], who agree on about 83% of epochs and least on N1 and on epochs around stage transitions [24]. Early deep networks learned stage features directly from one raw EEG channel [25]; later ones read a sequence of epochs around each epoch they label, as in SeqSleepNet [26], U-Sleep [23] and SleepTransformer [27], while open tools such as YASA [28] score a whole night from features of the full recording. Because performance drops when a network meets a cohort or montage outside its training data [29], recent models are trained across many datasets and tested on held-out ones [30]. Among open stagers, the Greifswald Sleep Stage Classifier (GSSC) [31] suits devices particularly well: it was trained across many electrode set-ups, including portable systems, and can run in real time inside brain–computer interfaces. Bringing such stagers onto devices has followed two lines: transferring networks from PSG to new sensors, such as textile headbands and ear-EEG [13,32,33], and adapting staging to real-time use. Along the second line, early systems staged each epoch as it arrived, on a server that also triggered acoustic stimulation [34] or with recurrent networks on home EEG [35]; recent work runs staging on embedded hardware [36], scores 5-s mini-epochs [37] or tests how electrode set-up, causal context and population shape online scoring for stimulation [21].

However, a stager that labels epochs well does not yet detect awakenings well. Its training objective does not target the event: awakenings after sleep onset are short and rare, a few bouts in a night dominated by sleep, and their first minutes are transitional epochs on which human scorers also agree least [24]. A network trained on stage labels sees few such examples, and on a cohort or sensor outside its training data, where staging performance already drops [29], these epochs grow harder still. Its usual evaluation does not show what a device needs either. Epoch accuracy and Cohen’s kappa are computed offline over the whole night, whereas a device needs to know how many awakenings it catches, how many false alarms it raises per hour of sleep and whether these numbers hold on cohorts and sensors it was not trained on. Brief events such as sleep spindles are evaluated by matching each detected event to a scored one [38]. An awakening detector needs the same event-level evaluation, run on past signal only.

We therefore developed WakeSentry, a real-time awakening detector, and evaluated it as a device would run it. WakeSentry pairs a streaming staging network with an awakening module that learns the first minutes of each wake bout, and trains the two on stage and awakening labels together, so that the network learns the moment of waking directly. We ran each detector on past signal only and counted the awakenings it caught at a matched false-alarm rate and with thresholds set in advance, holding each of five PSG cohorts out of training in turn and testing on two forehead headbands, one of them worn at home. Under this protocol, WakeSentry detected more awakenings than the published GSSC in every cohort and on both headbands, and on PSG more on average than U-Sleep and the offline stager YASA. On the headbands, the gain depended on the awakening module.

## 2. Materials and Methods

### 2.1. Datasets

We used five public PSG cohorts: the Sleep Cassette recordings of Sleep-EDF Expanded [39,40], the Haaglanden Medisch Centrum sleep staging database (HMC) [29], the training set of the PhysioNet/Computing in Cardiology Challenge 2018 [41], the PSG recordings of the Bitbrain Open Access Sleep dataset (BOAS) [42] and the CAP Sleep Database [43]. BOAS also contains a forehead EEG headband recorded in the same nights, which served as the first wearable test set. The second came from the open-access release of Wearanize+ [44,45], in which healthy adults aged 18 to 39 wore a Zmax forehead headband at home together with PSG. We used the 96 nights whose released file contained the right forehead channel of the Zmax headband, with the hypnogram of the first scorer as reference. Supplementary Methods S1 describes how we moved all Zmax decisions onto the clock of the hypnogram for each night. Sleep-EDF was recorded at home with an ambulatory recorder and scored following the Rechtschaffen and Kales manual, with stages 3 and 4 merged into N3 [39,40]. The PhysioNet 2018 recordings were made at the sleep laboratory of Massachusetts General Hospital and scored by clinical staff following the AASM manual [41]. In BOAS, every PSG epoch took the label given by at least two of three expert scorers [42]. For both headbands, the reference was the hypnogram scored on the simultaneous PSG. Every model used one EEG channel; in CAP it was F4-C4 or, where that was absent, the first available of F3-C3, F4-A1, F3-A2, C4-A1 and C3-A2; five of the 108 CAP recordings had none of these derivations or scoring that did not align with the signal and were left out. Table 1 gives the population, channel, persons and nights of each test set, with sex as recorded in each dataset.

**Table 1.** Test sets.

| Test set | Population | EEG channel | Nights | Persons | Age, y, median (IQR) | Female/male | Scored awakenings |
|---|---|---|---|---|---|---|---|
| Sleep-EDF | Healthy adults, recorded at home | Fpz-Cz | 153 | 78 | 57 (38–73) | 41/37 | 1,305 |
| HMC | Patients referred to a sleep clinic | F4-M1 | 151 | 151 | — | 66/85 | 1,168 |
| PhysioNet 2018 | Patients referred to a sleep laboratory | F4-M1 | 994 | 994 | 56 (46–66) | 328/666 | 7,286 |
| BOAS PSG | Adults without known sleep disorders | F4 | 128 | 100 | 31 (26–48) | 60/40 | 1,162 |
| CAP | Patients with sleep disorders and 16 controls | F4-C4* | 103 | 103 | 40 (28–61) | 40/60 | 407 |
| PSG total |  |  | 1,529 | 1,426 |  | 535/888 | 11,328 |
| BOAS headband | Same nights as BOAS PSG | Channel 1 | 128 | 100 | 31 (26–48) | 60/40 | 1,162 |
| Zmax headband | Healthy young adults, recorded at home | Right forehead | 96 | 96 | 22 (20–25) | 64/32 | 490 |
| Headband total |  |  | 224 | 196 |  | 124/72 | 1,652 |

Age: median (interquartile range) of the persons with recorded age; HMC gives no age per person. Female/male: sex as recorded in each dataset; HMC reports it for the whole cohort only, and CAP does not record it for 3 persons. \*CAP: F4-C4 or, where absent, the first available of F3-C3, F4-A1, F3-A2, C4-A1 and C3-A2. PhysioNet 2018: median apnoea–hypopnoea index 17.5 (IQR 8.8–25.9) scored respiratory events per hour of sleep. Zmax headband: home nights of Wearanize+, scored on the simultaneous PSG. Scored awakening: at least one wake epoch after 10 min of uninterrupted sleep.

### 2.2. Overview of WakeSentry

WakeSentry is a real-time awakening detector built from three parts: a convolutional encoder, a recurrent context network and an awakening module (Fig. 1). Every 3 s it takes the last 30 s of the frontal EEG, passes them through the frozen encoder module, updates the recurrent context network, and outputs two probabilities: that the person is in N2 or N3 sleep, and that an awakening is under way. A decision rule turns these probabilities into ‘asleep’ and ‘awake’ calls. Apart from about 0.1 s used by the resampling filter, only signal up to the current moment enters any step. One 3-s decision took a median of 9 ms on one thread of a desktop processor (Supplementary Methods S1).

**Fig. 1.**
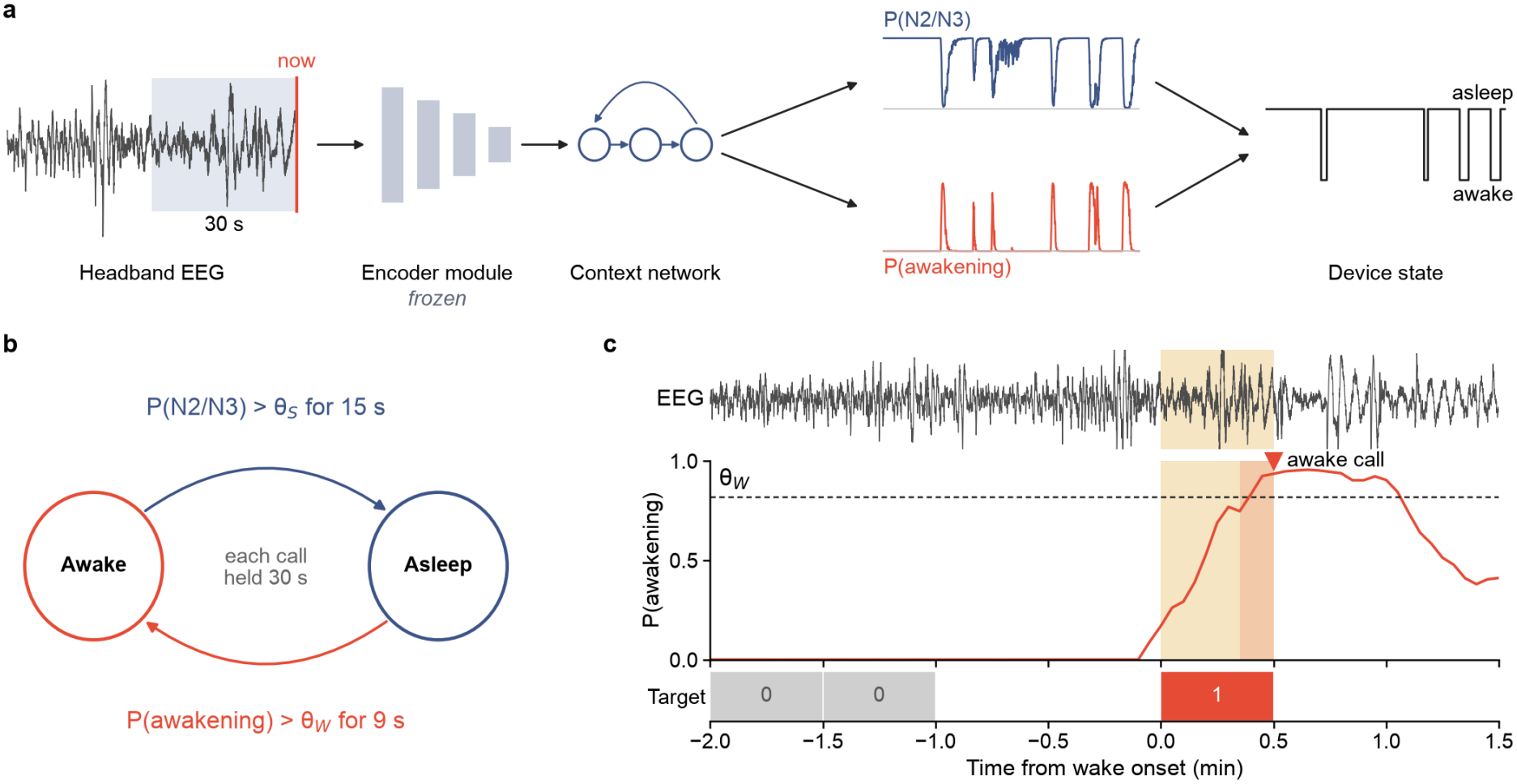
WakeSentry detects awakenings from one frontal EEG channel using only past signal. (a) Every 3 s, the last 30 s of EEG yield the probability of N2 or N3 sleep and, through the awakening module, the probability of an awakening; traces show 90 min of the headband night in Fig. 4a. The module’s training target is 1 for the first 2 min of wake, 0 for stable sleep and blank elsewhere. (b) The decision rule turns the two probabilities into ‘asleep’ and ‘awake’ calls at the thresholds θS and θW. (c) One scored awakening (yellow), called ‘awake’ 30 s after wake onset.

### 2.3. Streaming staging module

The encoder turns each 30-s epoch of EEG into a feature vector, and the context network, built of gated recurrent units, carries its state from epoch to epoch, so that each epoch is labelled from past signal alone. To obtain a decision every 3 s, we ran ten copies of the network side by side, offset by 3 s; at each step the copy whose epoch had just ended provided the output. The signal was band-pass filtered at 0.3–30 Hz with a causal Butterworth filter, resampled to 85.33 Hz and z-scored with the mean and standard deviation of the signal recorded so far.

### 2.4. Awakening module

Stage labels give a network few examples of the moment of waking, so we added a module trained on that moment. The module is a logistic unit on the 512 features of the context network. Its target is 1 for the first 2 min of each wake bout that follows sleep, 0 for stable sleep, defined as sleep with no wake epoch within 1 min, and is ignored elsewhere. Sleep epochs within 1 min of wake are left out because stage transitions are the epochs that scorers agree on least [24]. The loss is a weighted binary cross-entropy. The module’s probability, rather than the wake probability of the staging network, is the evidence for an ‘awake’ call.

### 2.5. Training

The encoder and context network started from the published weights of GSSC [31], which was trained on 2,652 PSG nights from SHHS, CFS, the Wisconsin Sleep Cohort and the Nationwide Children’s Hospital Sleep DataBank, none of which is among our test sets. Training followed a leave-one-dataset-out scheme: for each test cohort, the network was trained on the other four. For both headbands, we applied the model trained without BOAS, so that no head-band recording and no BOAS night entered training. In each training cohort, 15% of the persons, and at least two, were held out for early stopping and for choosing decision thresholds; the same persons were held out in every fold. Training had two steps: the module was first trained alone on the unchanged network with AdamW [46], and the module and all weights of the context network were then trained on the stage and module losses together, with the encoder frozen. The training configuration was settled during development on the five PSG cohorts and the BOAS headband, and the Zmax recordings were analysed after it was fixed. Supplementary Methods S1 gives the losses, learning rates, regularisation and stopping rule.

### 2.6. Decision rule and operating point

The device starts in the awake state. It calls ‘asleep’ when the N2+N3 probability has stayed above a threshold for 15 s, and ‘awake’ when the awakening probability has stayed above a second threshold for 9 s; after each call the state is held for 30 s. The dwell times stop a single noisy step from changing the state, and the hold time stops it from flickering near a threshold. The operating point, one threshold for each call, was chosen on the held-out persons of the training cohorts, never on the test cohort, and was used unchanged in the device analyses of Sections 3.4 and 3.5. Among threshold pairs with at most 0.5 false awakenings per hour of stable sleep and at most one false onset per hour of wake, we took the pair with the highest mean of onset hit rate and awakening sensitivity. The same procedure was applied to GSSC.

### 2.7. Comparison methods

We compared WakeSentry with three published stagers, each run on the same channel and nights. GSSC was run with its published weights through the streaming pipeline described above. U-Sleep [23] was run with the single-channel EEG weights released with SLEEPYLAND, which were trained on NSRR cohorts, none of which is among our test sets [47]. At each 3-s step it read the past 17.5 min of signal, as described in Supplementary Methods S1. YASA 0.8.1 [28] was run with its EEG-only model on the whole night at 100 Hz, as it was designed, and the probabilities of each epoch were assigned to the 3-s steps within it. YASA recommends a central derivation, so the frontal channel lay outside its preferred input. All three used the same decision rule and the same choice of operating points as WakeSentry.

### 2.8. Awakening events and measures

A scored awakening was a run of one or more wake epochs after at least 10 min of uninterrupted sleep. It counted as detected when an ‘awake’ call fell between 30 s before its start and 60 s after its end; the window opens before the first wake epoch because waking can begin within the preceding scored epoch. A false awakening was an ‘awake’ call during stable sleep, expressed per hour of stable sleep spent in the asleep state. A scored sleep onset was the first sleep epoch after lights-off or after at least 2 min of wake, followed by at least 9 min of sleep within 10 min. An onset counted as hit when the ‘asleep’ call came within 5 min of the first N2 epoch. The primary measure was awakening sensitivity at 0.5 false awakenings per hour of stable sleep, a budget of about three false calls in a night with 6 h of stable sleep. To read it, we kept the onset threshold at its operating point, swept the awakening threshold over 30 values on the test nights and interpolated sensitivity at 0.5 false awakenings per hour, so that all detectors were compared at the same false-alarm rate on the same nights.

### 2.9. Closed-loop replay and statistics

To see what the detectors would mean for a stimulator, we replayed each night and used the device state as the gate of a stimulator that acts only while the device reports sleep. We counted the minutes of scored wake after sleep onset during which the gate stayed open, and the share of scored N2 and N3 time during which it was open. Confidence intervals came from 2,000 bootstrap resamples of persons, paired across methods, so that nights of the same person stayed together. The averages over the PSG cohorts and over the two headbands gave each test set equal weight. The primary comparison was the PSG mean of WakeSentry minus GSSC; the other intervals are not adjusted for multiplicity. For the nightly wake after sleep onset (WASO) and number of awakenings reported by the device, we computed the mean bias, 95% limits of agreement and intraclass correlation against the hypnogram, on nights whose first scored sleep onset followed the wake at lights-off and on which the device also marked onset. Device WASO was the time in the awake state from the device’s sleep onset to the last scored sleep epoch; because the device returns to sleep only on an N2 or N3 call, N1 and REM after an awake call counted as wake. Sensitivity at the fixed thresholds was also compared by age, by sex, by apnoea–hypopnoea index in PhysioNet 2018, by diagnosis in CAP, by the length of each wake bout and by the stage scored before it. Finally, we shortened and lengthened each dwell time and the hold time in turn and chose the thresholds again on the training cohorts, as Supplementary Methods S1 describes.

## 3. Results

### 3.1. Participants and recordings

The five PSG cohorts contributed 1,529 test nights from 1,426 persons. These nights contained 11,328 scored awakenings after sleep onset, and Table 1 gives the counts for each cohort. The 128 BOAS nights were also recorded with a headband and contained 1,162 awakenings. The Zmax headband added 96 home nights, one per person, with 490 awakenings. Women made up 38% of the PSG persons and 63% of the headband persons. Fig. 2a shows which cohorts trained the model applied to each test set. The median number of scored awakenings per night ranged from 3 in CAP to 9 in BOAS, and Fig. 2b,c gives the distributions of awakenings and WASO.

**Fig. 2.**
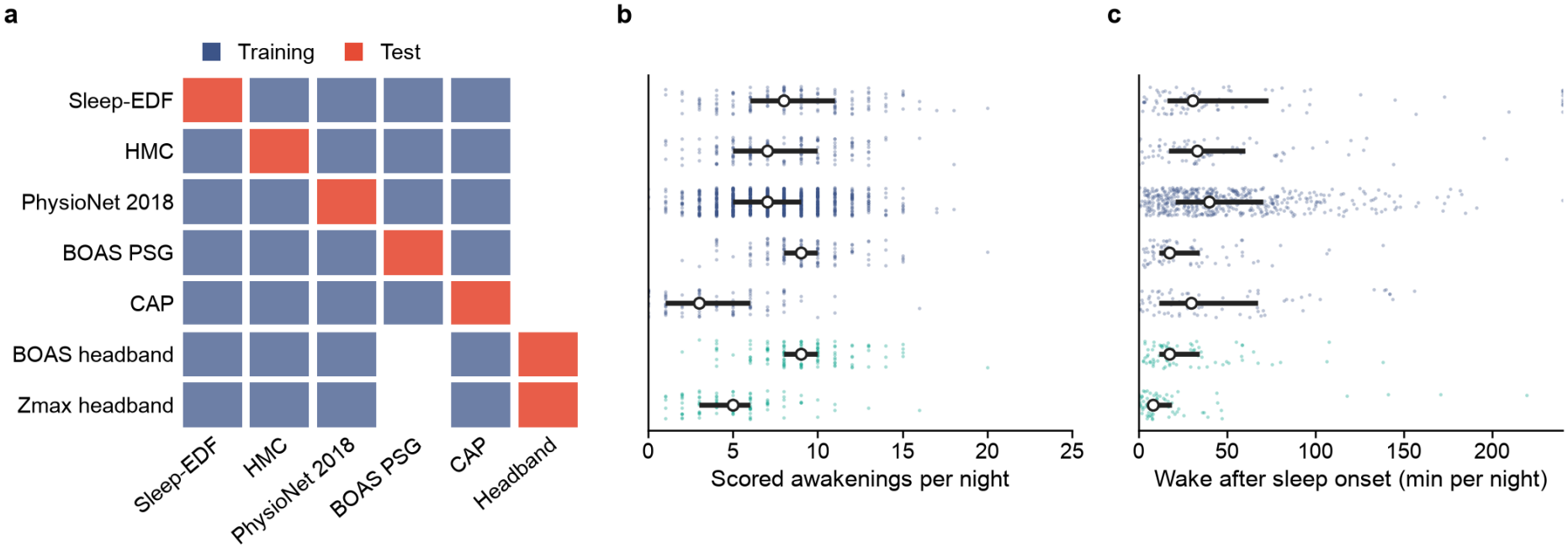
Each test set was held out of training, and the test sets differ in how often people wake. (a) Training data (blue) and test data (red) for each test set; both headbands use the model trained without BOAS. (b) Scored awakenings and (c) wake after sleep onset per night, the latter on the nights of the night-level analysis; dots are nights, white circles medians and bars interquartile ranges.

### 3.3. Awakening detection on polysomnography

To test whether the awakening module and retraining improve real-time detection, we compared WakeSentry with the published GSSC at 0.5 false awakenings per hour of stable sleep. WakeSentry had higher sensitivity in all five PSG cohorts, with gains from 3.3 percentage points on Sleep-EDF to 10.0 points on HMC, as Table 2 and Fig. 3b show. In Fig. 3a, the WakeSentry curve lay above that of GSSC from 0.1 to 2 false awakenings per hour in HMC and CAP, and over more than half of this range in the other three cohorts. Averaged with equal weight over the cohorts, sensitivity rose from 63.8% to 70.5%. The gain of 6.7 points had a 95% confidence interval of 5.4 to 7.9 points, and every cohort’s interval lay above zero. At the same false-alarm cost, WakeSentry caught more awakenings in healthy sleepers and in patients alike.

**Table 2.**
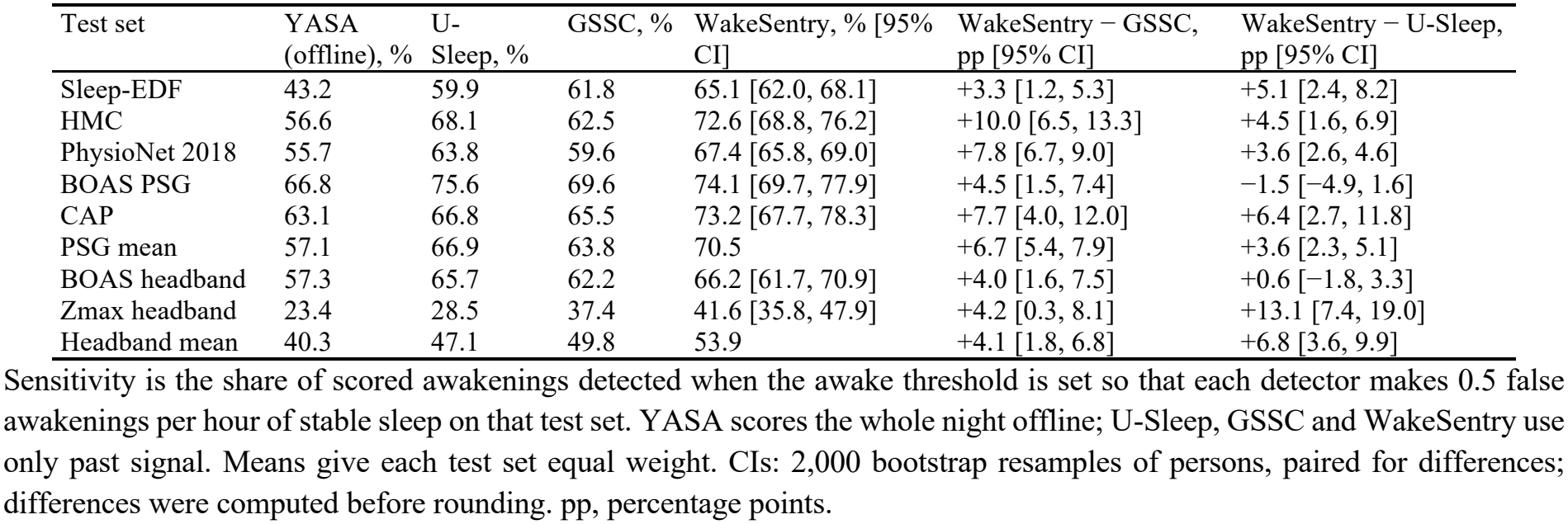
Awakening sensitivity of four detectors at 0.5 false awakenings per hour.

| Test set | YASA (offline), % | U-Sleep, % | GSSC, % | WakeSentry, % [95% CI] | WakeSentry − GSSC, pp [95% CI] | WakeSentry − U-Sleep, pp [95% CI] |
|---|---|---|---|---|---|---|
| Sleep-EDF | 43.2 | 59.9 | 61.8 | 65.1 [62.0, 68.1] | +3.3 [1.2, 5.3] | +5.1 [2.4, 8.2] |
| HMC | 56.6 | 68.1 | 62.5 | 72.6 [68.8, 76.2] | +10.0 [6.5, 13.3] | +4.5 [1.6, 6.9] |
| PhysioNet 2018 | 55.7 | 63.8 | 59.6 | 67.4 [65.8, 69.0] | +7.8 [6.7, 9.0] | +3.6 [2.6, 4.6] |
| BOAS PSG | 66.8 | 75.6 | 69.6 | 74.1 [69.7, 77.9] | +4.5 [1.5, 7.4] | −1.5 [−4.9, 1.6] |
| CAP | 63.1 | 66.8 | 65.5 | 73.2 [67.7, 78.3] | +7.7 [4.0, 12.0] | +6.4 [2.7, 11.8] |
| PSG mean | 57.1 | 66.9 | 63.8 | 70.5 | +6.7 [5.4, 7.9] | +3.6 [2.3, 5.1] |
| BOAS headband | 57.3 | 65.7 | 62.2 | 66.2 [61.7, 70.9] | +4.0 [1.6, 7.5] | +0.6 [−1.8, 3.3] |
| Zmax headband | 23.4 | 28.5 | 37.4 | 41.6 [35.8, 47.9] | +4.2 [0.3, 8.1] | +13.1 [7.4, 19.0] |
| Headband mean | 40.3 | 47.1 | 49.8 | 53.9 | +4.1 [1.8, 6.8] | +6.8 [3.6, 9.9] |

Sensitivity is the share of scored awakenings detected when the awake threshold is set so that each detector makes 0.5 false awakenings per hour of stable sleep on that test set. YASA scores the whole night offline; U-Sleep, GSSC and WakeSentry use only past signal. Means give each test set equal weight. CIs: 2,000 bootstrap resamples of persons, paired for differences; differences were computed before rounding. pp, percentage points.

**Fig. 3.**
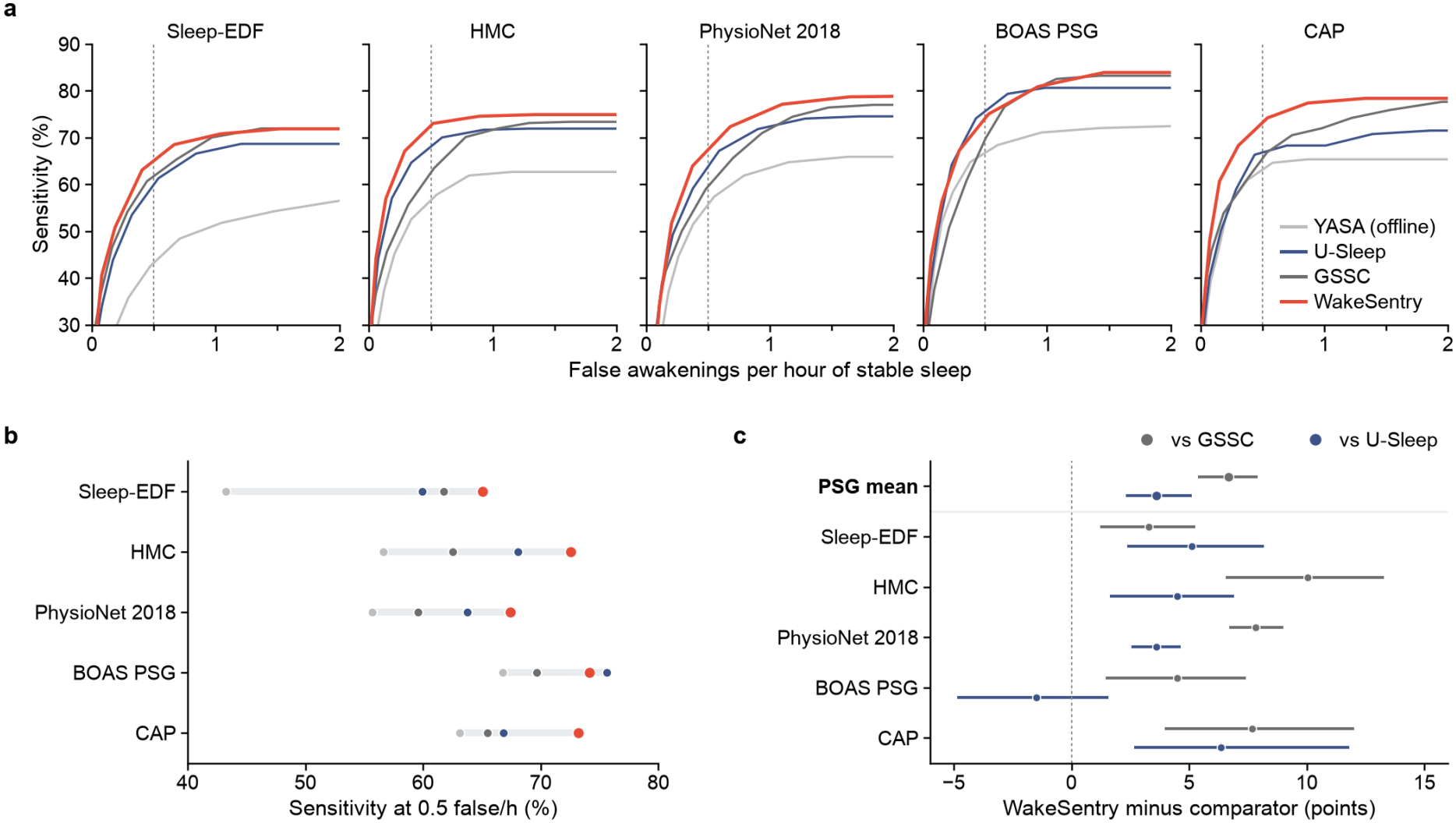
On PSG, WakeSentry detects more awakenings than GSSC in every cohort and, on average, more than U-Sleep and YASA. (a) Sensitivity against false awakenings per hour of stable sleep; the dashed line marks 0.5 per hour. (b) Sensitivity at 0.5 false awakenings per hour. (c) WakeSentry minus GSSC (grey) and minus U-Sleep (blue), with 95% person-bootstrap intervals; the top row is the mean of the five cohorts.

To place this gain among published stagers, we ran U-Sleep and YASA on the same nights and channels. Averaged over the PSG cohorts, WakeSentry exceeded U-Sleep by 3.6 percentage points (95% CI 2.3–5.1), with intervals above zero in four of the five cohorts. Its margin over YASA was 13.4 points, and Fig. 3c gives the paired differences for every cohort. U-Sleep, run as published, was itself 3.1 points more sensitive than GSSC on PSG, so WakeSentry turned the weaker network into the stronger.

### 3.3. Awakening detection on forehead headbands

To test whether the gain holds on wearable sensors, we applied the models trained without BOAS to two forehead headbands: the BOAS headband, recorded in the same nights as BOAS PSG, and the Zmax headband, worn at home by young adults [44]. WakeSentry detected more awakenings than GSSC on both, by 4.0 points on BOAS and 4.2 points on Zmax. The mean gain had a 95% confidence interval of 1.8 to 6.8 points. Over the two headbands it also exceeded U-Sleep by 6.8 points, almost entirely on the home recordings: there all four detectors found fewer awakenings and WakeSentry led U-Sleep by 13.1 points, whereas on the BOAS headband the two were level. Fig. 4a–c shows one headband night, the curves and the paired differences. On both headbands, the WakeSentry curve lay above that of GSSC at every false-alarm rate from 0.1 to 2 per hour. The gain carried over to forehead sensors unseen in training.

**Fig. 4.**
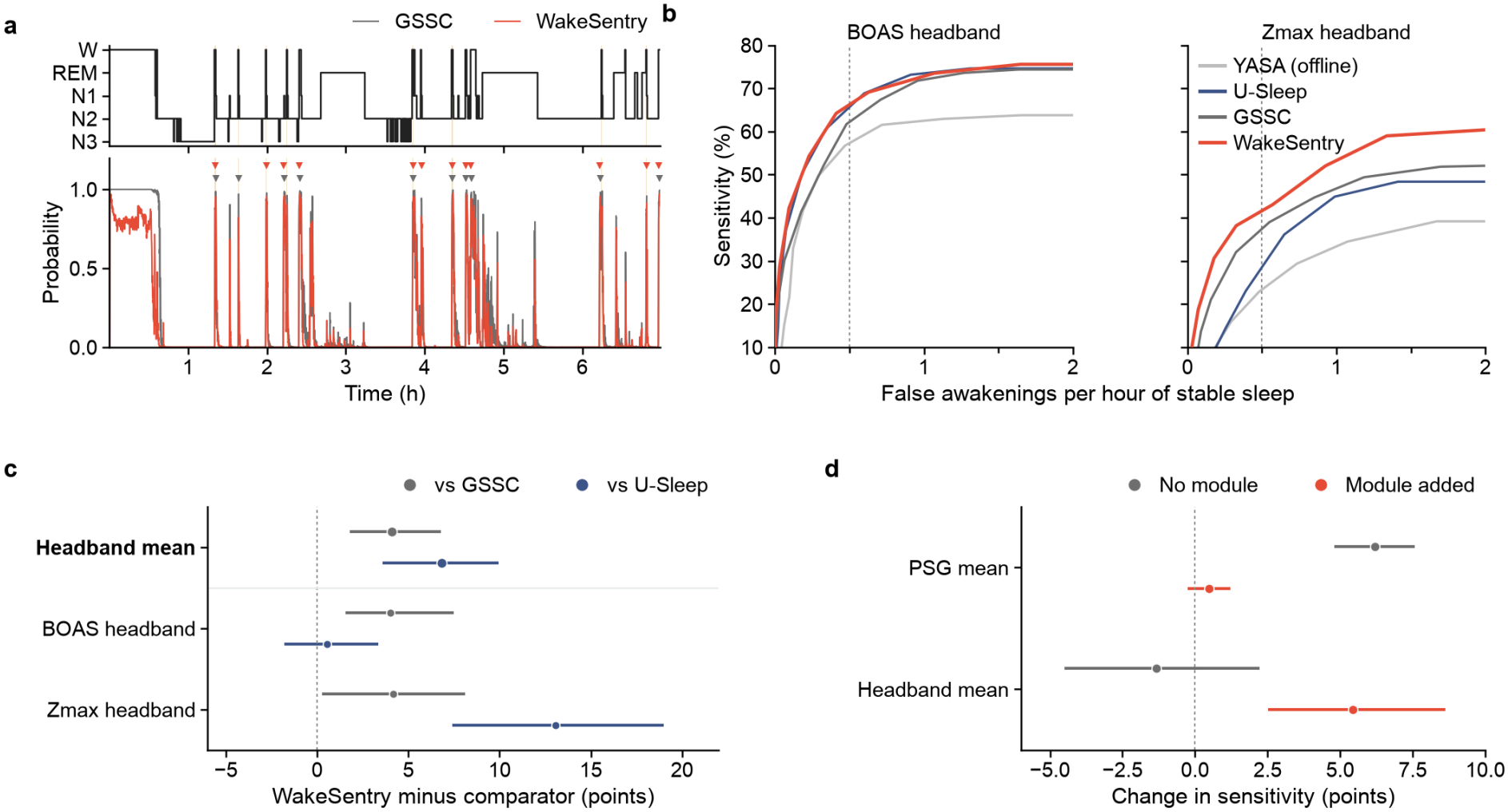
On two forehead headbands never seen in training, WakeSentry keeps its advantage, which is lost without the awakening module. (a) One BOAS headband night: hypnogram, GSSC wake probability and WakeSentry awakening probability; triangles mark awake calls and yellow bands scored wake. (b) Sensitivity against false awakenings per hour. (c) WakeSentry minus GSSC (grey) and minus U-Sleep (blue) at 0.5 false awakenings per hour, with 95% intervals; the top row is the mean of both headbands. (d) Change in sensitivity from training without the module (grey, versus GSSC) and from adding the module (red).

### 3.4. Detection with thresholds fixed in advance

To see how the detectors behave as devices, we ran them at the thresholds set in advance on the training cohorts. Fig. 5a and Supplementary Table S1 give the results. WakeSentry detected more awakenings than GSSC in six of the seven test sets, by 5.5 to 17.1 percentage points, and each of these six intervals excluded zero. Its false-awakening rate ranged from 0.15 to 0.40 per hour, and it was higher than that of GSSC on Sleep-EDF and both headbands, the three test sets with the largest gains in sensitivity. GSSC exceeded the budget of 0.5 per hour only on Physio-Net 2018, where WakeSentry halved the false awakenings at a sensitivity about 2 points lower. With shorter or longer dwell and hold times, WakeSentry still detected more awakenings than GSSC in at least six test sets, as Supplementary Table S5 shows. The median delay from the start of a wake bout to its detection was 30 to 41 s for WakeSentry and 27 to 49 s for GSSC. WakeSentry called most detected awakenings within the first minute of wake: 85% to 95% in every test set. Fig. 5b shows that this share was larger than that of GSSC in six of the seven test sets, with intervals above zero, and level on PhysioNet 2018. A device can therefore run with thresholds fixed before use.

**Fig. 5.**
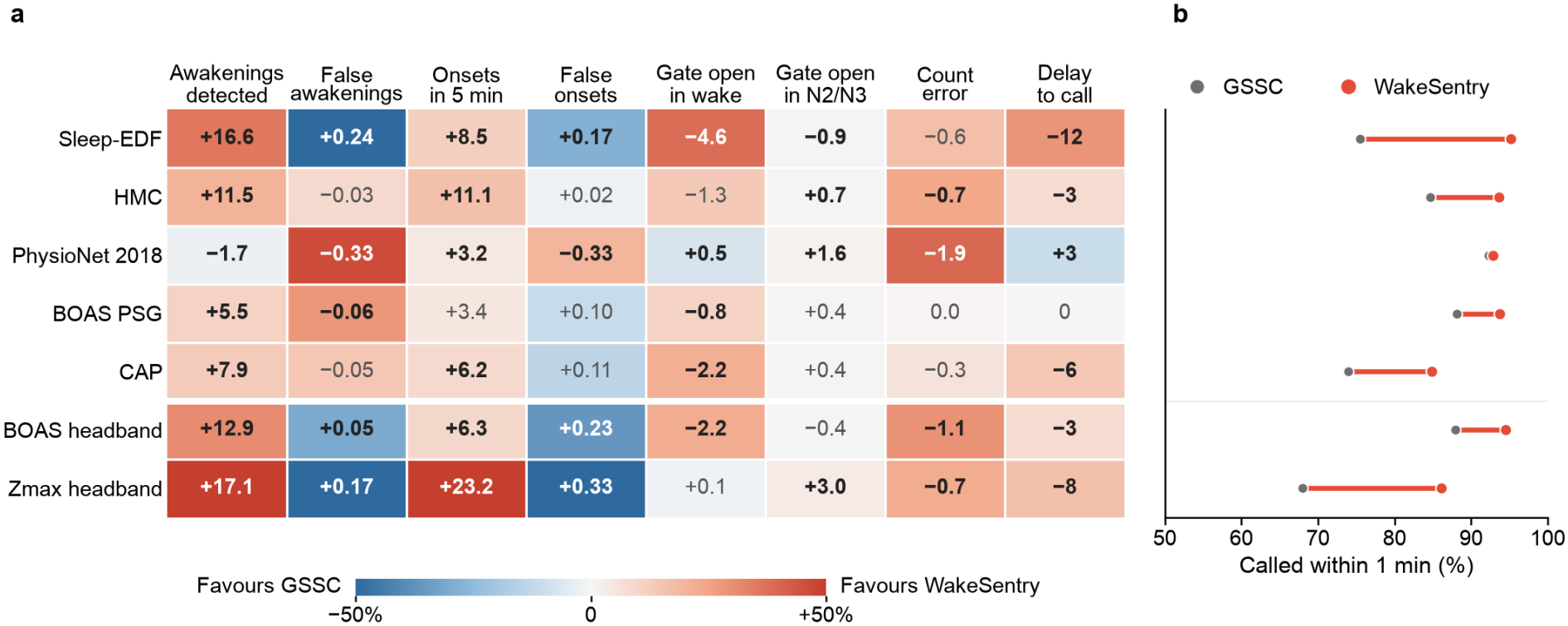
Run as a device, WakeSentry detects more awakenings than GSSC in six of seven test sets and calls them sooner. Both detectors use the operating points fixed on the training cohorts. (a) Change from GSSC to WakeSentry in each measure (Sections 2.8 and 2.9) and test set, in percentage points, events per hour, minutes per night, awakenings per night or seconds; colour gives the change relative to GSSC, red favouring WakeSentry and blue favouring GSSC, and bold values have 95% person-bootstrap intervals that exclude zero, without adjustment for multiplicity. (b) Share of detected awakenings called within 1 min of wake onset, GSSC (grey) and WakeSentry (red); red lines mark differences whose intervals lie above zero.

At the same operating point, WakeSentry called sleep onset within 5 min of the first N2 epoch more often than GSSC in all seven test sets, with intervals above zero in six. False onset calls rose in six test sets, with intervals above zero on Sleep-EDF and both headbands, and fell by 0.33 per hour on PhysioNet 2018, the largest cohort. A stimulator can thus start closer to the true onset of sleep.

### 3.5. Closed-loop replay, night-level measures and subgroups

To translate detection into device terms, we replayed each night with the device state gating a stimulator. With GSSC as the gate, the stimulator stayed open for 7.2 to 12.0 min of scored wake per night. With WakeSentry, this time fell in five of the seven test sets, by 0.8 to 4.6 min per night, with intervals excluding zero in four. It rose on PhysioNet 2018 and Zmax, by 0.5 and 0.1 min. The WakeSentry gate stayed open for at least 85% of scored N2 and N3 time in every test set and never fell more than 1 point below GSSC. A WakeSentry gate thus spends less stimulation on wake in most test sets.

Night-level measures used the nights on which the scorers and the device both marked a first sleep onset after lights-off, 48% to 72% of each test set. On these nights, the number of awakenings that WakeSentry reported came closer to the scored number than that of GSSC on HMC, PhysioNet 2018 and both headbands, and matched GSSC on the other three test sets. Its nightly count also had a higher intraclass correlation with the scored count than that of GSSC in six of the seven test sets. Fig. 5a includes the change in count error, and Supplementary Table S2 gives the limits of agreement. The nightly WASO summed from the calls ran above the scored WASO for both detectors, by a mean of 22 to 41 min for WakeSentry, and further above it than that of GSSC in six of the seven test sets. The device thus reports the nightly awakening count well.

At the thresholds set in advance, WakeSentry detected more awakenings than GSSC in persons younger than 40 and in those aged 60 or older, by 5.0 and 3.4 points. Between 40 and 59 it detected 1.6 points fewer. Within PhysioNet 2018, it detected fewer awakenings than GSSC in three of four bands of apnoea–hypopnoea index, by up to 3.8 points, at a lower false-awakening rate than GSSC in every band. Among CAP patients, gains were largest in nocturnal frontal lobe epilepsy and REM sleep behaviour disorder, by 7.6 and 9.6 points. Sensitivity was unchanged in insomnia and 4.3 points lower in periodic leg movements. By sex, the gain was 2.9 points in women and 1.2 in men. Supplementary Table S3 lists all subgroups. The smaller gains came mainly from PhysioNet 2018, where WakeSentry raised fewer false alarms.

To see which awakenings were missed, we split sensitivity by the length of each wake bout and by the stage before waking, as Supplementary Table S4 shows. Single-epoch bouts made up nearly half of the scored awakenings on PSG and were missed most often by both detectors. WakeSentry caught more bouts of two or three epochs than GSSC in every test set, by 5.9 to 25.2 points, and more awakenings from N3, by 4.5 to 38.5 points. Only in these two groups did WakeSentry gain in every test set.

### 3.6. Contribution of the awakening module

To isolate the awakening module, we removed it and trained the context network on stage labels alone, with the same data, held-out persons and stopping rule. On the two headbands, removing the module lowered sensitivity by 5.4 points on average (95% CI 2.5–8.6), and the network trained without it gained nothing over GSSC. The loss was 4.8 points on the BOAS headband and 6.0 points on Zmax. On PSG, removing the module lowered sensitivity by half a point, a difference whose interval included zero, and Fig. 4d sets both changes side by side. So on the headbands the gain came from the module, and on PSG from retraining.

## 4. Discussion

We asked whether awakenings can be detected in real time from one frontal EEG channel across cohorts and sensors. WakeSentry detected more awakenings than the published GSSC at a matched false-alarm rate in all five PSG cohorts and on two forehead headbands, while running causally with one decision every 3 s. On the headbands, this gain depended on the awakening module. On PSG it also caught more awakenings on average than U-Sleep run on past signal and YASA run on the whole night.

### 4.1. Implications for closed-loop and home devices

Within a budget of 0.5 false awakenings per hour of stable sleep, WakeSentry missed on PSG about one fifth fewer awakenings than GSSC. The closed-loop replay shows what this means for a stimulator: in five of seven test sets, a stimulator gated by WakeSentry stayed open during wake for fewer minutes. Older adults wake more often during the night [2,3] and are a main target group of slow-oscillation stimulation [15,48], so a gate that closes at awakenings matters most for them. In every test set, it also caught more awakenings from N3 than GSSC, and N3 is the stage in which slow-oscillation stimulation delivers its sounds. This matters because cues delivered in wake do not strengthen memory [20], and sounds delivered to a waking sleeper add to the fragmentation that stimulation should avoid [1].

On the Zmax headband, WakeSentry detected more awakenings than GSSC and U-Sleep at the same false-alarm rate, and its nightly count came closer to the scored count, with a mean absolute error of 2.3 against 2.9 awakenings per night.

### 4.2. Comparison with previous work

These results complement earlier work on bringing staging to devices. Transfer from PSG to headbands and ear-EEG has been judged by epoch accuracy, which reached 82% on a textile headband [13] and 70% to 74% on in-ear EEG in older adults [33]. Real-time staging for stimulation has so far been judged on finding the state to stimulate: a dry-EEG headband detected N3 in real time with a sensitivity of 0.70 [16], and a consumer closed-loop system detected slow-wave sleep with 74% sensitivity [17], and algorithms that time sounds to slow-wave phase have been benchmarked on wearable EEG [49]. Among published stagers, U-Sleep holds up well across cohorts under a common evaluation [23,47], and on PSG it was more sensitive to awakenings than GSSC, though less than WakeSentry. The PhysioNet 2018 Challenge asked for arousals to be detected from whole recordings [41], whereas we detected online the longer awakenings that a device acts on. YASA, which reads the whole night [28], found fewer awakenings than each causal detector on every test set. This event-level view, long used for spindles [38], measures the step at which a closed loop stops stimulation, which epoch-level measures do not.

Two alternative explanations can be checked. Threshold choice does not explain the gain at the matched rate, because every detector was read at the same false-alarm rate on the same nights, with the onset threshold it kept from its training cohorts. A cohort-specific effect does not explain the gain either, because each cohort was left out of training in turn, the gain over GSSC appeared in all five, and it held on the home headband, which was analysed only after the configuration was fixed.

### 4.3. Why the awakening module matters on headbands

Waking is hard to see in one frontal channel: forehead EEG recognises N1 poorly because it lacks occipital electrodes [9], and the epochs around a transition between wake and sleep are those that scorers agree on least [24]. A module trained on the first minutes of wake gives the network direct examples of exactly these epochs. The ablation places its effect on the headbands. This pattern matches the finding that training a new head first and then the whole network preserves pretrained features under distribution shift, whereas training the whole network at once can distort them [50]. Because the module also brings this head-first schedule, its effect on the headbands combines the awakening target and the schedule. A headband differs from PSG in electrode position and signal quality, and so presents this kind of shift.

### 4.4. Strengths and limitations

The study has three strengths: each cohort and both headbands were held out of training, all detectors were compared on the same nights and false-alarm rate, and the test sets covered healthy sleepers, clinic patients and home recordings.

Three features of the study also set its scope. The headband results come from two forehead devices, and the home recordings had one night per person, so sensors at other sites, such as inear EEG, and multi-night home use are the next tests. The device returns to sleep only on an N2 or N3 call, so its summed WASO counted N1 and REM after an awakening as wake and ran above the scored value. Finally, the closed-loop results come from replayed nights, and the effect on stimulation outcomes will be measured in a live session.

Next, we will run WakeSentry inside a closed-loop auditory stimulation session, where it pauses stimulation at each detected awakening, and test it on multi-night home recordings, including people with insomnia, whose WASO varies widely between nights [5]. Because the awakening module reads only the features of a context network, we will also attach it to other sequence stagers such as U-Sleep and SeqSleepNet [26]. Finally, we will build a nightly WASO estimate that counts N1 and REM as sleep.

## 5. Conclusion

WakeSentry, with an awakening module trained on the moment of waking, detects awakenings in real time from one frontal EEG channel and past signal. At a matched false-alarm rate, it outperformed the published GSSC in healthy sleepers, in patients and on two forehead headbands unseen in training. With thresholds set in advance on the training cohorts, it kept false awakenings at or below 0.40 per hour in every test set, so developers of closed-loop stimulators and home EEG devices can fix its operating point before use.

## Data Availability

All recordings are publicly available: Sleep-EDF Expanded, the PhysioNet/CinC Challenge 2018 data, HMC and the CAP Sleep Database on PhysioNet, BOAS on OpenNeuro (ds005555) and Wearanize+ OA on the Radboud Data Repository (https://doi.org/10.34973/xrmf-5726). The code for all analyses and figures will be made publicly available upon journal publication. The trained WakeSentry weights and the operating thresholds will be released at the same time.

## CRediT authorship contribution statement

Han Cui: Conceptualization, Methodology, Software, Validation, Formal analysis, Investigation, Data curation, Visualization, Writing – original draft. Yuge Zhu: Validation, Writing – review & editing. Bing Li: Supervision, Writing – review & editing. Shaojia Huang: Supervision, Funding acquisition, Writing – review & editing. Tao Xu: Supervision, Project administration, Resources, Funding acquisition, Writing – review & editing.

## Ethics statement

This study analysed only de-identified recordings from public databases [29,39–45]. Each dataset was collected by its original investigators under the ethics approval and consent procedures described in the cited publications and is shared without information that identifies the participants. No new data were collected, the authors had no access to identifying information, and no attempt was made to re-identify any participant. Under the Measures for Ethical Review of Life Sciences and Medical Research Involving Humans of China (2023), research that uses lawfully obtained public data or anonymised information can be exempted from ethics review, and no further approval was therefore sought.

## Data availability

All recordings are publicly available: Sleep-EDF Expanded, the PhysioNet/CinC Challenge 2018 data, HMC and the CAP Sleep Database on PhysioNet, BOAS on OpenNeuro (ds005555) and Wearanize+ OA on the Radboud Data Repository (https://doi.org/10.34973/xrmf-5726). The U-Sleep weights are distributed with SLEEPYLAND [47]. The code for all analyses and figures will be made publicly available upon journal publication. The trained WakeSentry weights and the operating thresholds will be released at the same time.

## Declaration of generative AI and AI-assisted technologies in the manuscript preparation process

During the preparation of this work the authors used Claude (Anthropic) to assist with writing and reviewing the analysis code. After using this tool, the authors reviewed and edited the content as needed and take full responsibility for the content of the published article.

