## Supplementary Material for "WakeSentry: real-time awakening detection from single-channel EEG across sleep cohorts and forehead headbands"

### *Supplementary Methods S1*

**Training.** In the first step, the module alone was trained with AdamW at a learning rate of $10^{-3}$. In the second step, the module and all weights of the context network were trained together at a learning rate of $10^{-4}$, while the encoder stayed frozen. The class weights of the stage loss were inversely proportional to the square root of class frequency. Each step used 32 one-hour crops; training stopped when the validation stage loss, checked every 300 iterations, had not improved five times in a row. The awakening module saw, at each 3-s step, the share of its 30-s window that fell in positive or negative epochs, with the positive class weighted by the square root of the ratio of negative to positive steps. The stage loss was a class-weighted cross-entropy against the stages of the epochs that each 30-s window overlaps. AdamW used a weight decay of 0.01, gradients were clipped at a norm of 1.0, each step ran for at most 8,000 iterations, and the module loss had the same weight as the stage loss. In the second step, the starting network was kept if no fine-tuned version reached a lower validation loss.

**U-Sleep.** We reimplemented the network in PyTorch 2.15.0, and its outputs matched the original within $3 \times 10^{-6}$. The 17.5 min of signal ending at each decision were resampled to 128 Hz and robustly scaled within that window, and we took the wake probability and the summed N2 and N3 probabilities of the last epoch.

**Runtime.** One decision comprises the band-pass filter and resampling of the newest 3 s, z-scoring, the GSSC encoder and one step of the context network with the module. We timed 2,000 decisions on one thread of an AMD Ryzen 9 9950X3D desktop processor (AMD, Santa Clara, CA, USA), with PyTorch 2.15.0 in 32-bit floating point. A decision took a median of 9.1 ms, and 95% of decisions took under 9.7 ms. The network has 16.6 million parameters, 10.7 million of them in the encoder.

**Headband timing.** In BOAS, the headband and PSG files of each night have the same length. For each night, we computed the envelope of 1–4 Hz power in 1-s bins for the first headband channel and for PSG F4, and correlated the two envelopes over lags of up to 10 min. The correlation peaked at zero lag on 126 of 128 nights, and a search in 0.25-s steps kept each of these peaks at zero. On the other two nights, the two headband channels did not correlate with each other either, so their signal was too poor for this test.

**Zmax timing.** Wearanize+ stores the Zmax and PSG signals of each night in one file. The same envelope test, run on Zmax and PSG F4:A1, showed that the Zmax signals trailed the PSG by a median of 27 s. The movement signals of the two devices gave the same lag, within 3 s, on 83 nights. We used the EEG lag on these nights and wherever its peak exceeded the zero-lag correlation by at least 0.05. Otherwise we used the movement lag where its correlation reached 0.1, and the median lag on the remaining three nights. On the 60 nights with a clear peak in each half of the night, the lags of the two halves differed by at most 3 s. We then compared the hypnogram of the first scorer with the automatic U-Sleep scoring released with the dataset [23,24] at shifts of whole epochs.

The hypnogram matched the U-Sleep timing on 33 nights and sat one epoch later on 55. The remaining eight nights showed other whole-epoch shifts. The correlation of PSG delta power with scored N3 placed the hypnogram at the same position on 75 of 94 nights. The U-Sleep scoring served only to time the hypnogram, and the hypnogram of the first scorer remained the reference. We moved every decision by the Zmax lag minus the hypnogram offset, and ran YASA on the Zmax signal trimmed by the same amount. In the PSG cohorts, the same comparison against YASA staging of the detection channel found a whole-epoch shift that improved agreement by more than 1 point on at most 4% of nights.

**Second hypnogram.** Wearanize+ also releases a second PSG hypnogram. Read with our awakening definition, it marked wake within the detection window of 97% of the reference awakenings. It marked 1,112 awakenings in all, against 490 in the reference. The reference awakenings were thus present in both hypnograms, whereas the number per night depended on the hypnogram.

**Missed awakenings and decision rule.** Each scored awakening was assigned to a length class (one epoch, two or three epochs, four or more) and to the stage of the epoch before it, and detection used the window of the main analysis. For the decision rule, the dwell time for 'asleep' was set to 9 or 21 s instead of 15 s, that for 'awake' to 6 or 12 s instead of 9 s, and the hold time to 15 or 60 s instead of 30 s, one at a time. For every setting, the operating points of GSSC and WakeSentry were chosen again on the held-out persons of the training cohorts by the rule of Section 2.6.

**Table S1.** Both methods run as a device at the operating point fixed on the training cohorts.

| Measure | Method | Sleep-EDF | HMC | PhysioNet 2018 | BOAS PSG | CAP | BOAS headband | Zmax headband |
|---|---|---|---|---|---|---|---|---|
| **Awakening sensitivity, %** | GSSC | 46.5 | 55.7 | 65.7 | 50.8 | 60.4 | 41.4 | 21.0 |
| | WakeSentry | 63.1 | 67.1 | 63.9 | 56.3 | 68.3 | 54.3 | 38.2 |
| | Difference [95% CI] | +16.6 [13.3, 19.7] | +11.5 [8.5, 14.4] | −1.7 [−2.8, −0.6] | +5.5 [1.9, 9.3] | +7.9 [4.0, 11.6] | +12.9 [9.2, 17.0] | +17.1 [13.5, 20.9] |
| **False awakenings per hour of stable sleep** | GSSC | 0.16 | 0.32 | 0.70 | 0.21 | 0.36 | 0.18 | 0.16 |
| | WakeSentry | 0.40 | 0.29 | 0.37 | 0.15 | 0.31 | 0.23 | 0.33 |
| | Difference [95% CI] | +0.24 [0.19, 0.30] | −0.03 [−0.09, 0.03] | −0.33 [−0.36, −0.30] | −0.06 [−0.10, −0.02] | −0.05 [−0.12, 0.01] | +0.05 [0.01, 0.10] | +0.17 [0.12, 0.22] |
| **Onsets detected within 5 min, %** | GSSC | 71.5 | 67.6 | 71.4 | 71.8 | 69.4 | 65.5 | 27.4 |
| | WakeSentry | 80.0 | 78.7 | 74.7 | 75.2 | 75.6 | 71.8 | 50.5 |
| | Difference [95% CI] | +8.5 [4.7, 12.6] | +11.1 [7.4, 14.8] | +3.2 [1.9, 4.6] | +3.4 [−0.3, 7.6] | +6.2 [1.2, 11.0] | +6.3 [1.5, 12.0] | +23.2 [16.0, 30.9] |
| **False onsets per hour of wake** | GSSC | 0.61 | 1.03 | 1.64 | 0.89 | 0.79 | 0.62 | 0.31 |
| | WakeSentry | 0.78 | 1.05 | 1.31 | 0.99 | 0.90 | 0.85 | 0.64 |
| | Difference [95% CI] | +0.17 [0.06, 0.28] | +0.02 [−0.15, 0.18] | −0.33 [−0.41, −0.25] | +0.10 [−0.07, 0.26] | +0.11 [−0.03, 0.26] | +0.23 [0.09, 0.37] | +0.33 [0.11, 0.56] |
| **Gate open during wake, min per night** | GSSC | 12.0 | 9.1 | 7.2 | 7.6 | 10.1 | 9.7 | 8.4 |
| | WakeSentry | 7.4 | 7.9 | 7.8 | 6.8 | 7.9 | 7.5 | 8.6 |
| | Difference [95% CI] | −4.6 [−5.7, −3.7] | −1.3 [−2.5, 0.1] | +0.5 [0.2, 0.9] | −0.8 [−1.4, −0.3] | −2.2 [−3.3, −1.1] | −2.2 [−3.4, −1.2] | +0.1 [−2.1, 3.3] |
| **Gate open during N2+N3, %** | GSSC | 97.3 | 93.4 | 95.0 | 93.1 | 94.8 | 91.4 | 82.9 |
| | WakeSentry | 96.4 | 94.1 | 96.5 | 93.5 | 95.2 | 91.0 | 86.0 |
| | Difference [95% CI] | −0.9 [−1.6, −0.4] | +0.7 [0.0, 1.5] | +1.6 [1.4, 1.8] | +0.4 [−0.1, 0.9] | +0.4 [−0.1, 0.9] | −0.4 [−1.5, 0.6] | +3.0 [0.8, 5.3] |

| | | | | | | | | |
|---|---|---|---|---|---|---|---|---|
| **Awakening-count error (absolute)** | GSSC | 3.3 | 2.9 | 4.9 | 3.0 | 4.3 | 3.7 | 2.9 |
| | WakeSentry | 2.7 | 2.2 | 3.0 | 3.0 | 4.1 | 2.6 | 2.3 |
| | Difference [95% CI] | −0.6 [−1.4, 0.3] | −0.7 [−1.2, −0.2] | −1.9 [−2.3, −1.6] | 0.0 [−0.6, 0.5] | −0.3 [−0.9, 0.4] | −1.1 [−1.7, −0.5] | −0.7 [−1.1, −0.2] |
| **Median delay to awake call, s** | GSSC | 42 | 36 | 27 | 33 | 42 | 36 | 49 |
| | WakeSentry | 30 | 33 | 30 | 33 | 36 | 33 | 41 |
| | Difference [95% CI] | −12 [−15, −9] | −3 [−9, −3] | +3 [3, 6] | 0 [−3, 0] | −6 [−9, −3] | −3 [−6, −3] | −8 [−15, −6] |

Both methods run at the thresholds chosen on the validation persons of the training cohorts (at most 0.5 false awakenings and 1 false onset per hour) and then fixed for the test set. Gate: a stimulator that is allowed to fire only while the device reports sleep. Awakening-count error: absolute difference between the device estimate and the scored value per night. Delay: from the start of a detected awakening to the awake call; decisions fall on a 3-s grid, so the percentile intervals of the median delay can share a bound with the estimate. Awakening-count error: nights used for night-level agreement (Table S2). CIs: 2,000 paired bootstrap resamples of persons. Differences were computed before rounding.

**Table S2.** Agreement of the nightly WASO and number of awakenings with the hypnogram.

| Test set | Nights | Measure | GSSC bias [95% LoA] | GSSC ICC | WakeSentry bias [95% LoA] | WakeSentry ICC | Difference in MAE [95% CI] |
|---|---|---|---|---|---|---|---|
| Sleep-EDF | 87 of 153 | WASO, min | +17.7 [−54.5, 89.9] | 0.95 | +40.2 [−42.3, 122.6] | 0.91 | +19.5 [14.0, 25.4] |
| | | Awakenings per night | −1.9 [−9.5, 5.7] | 0.44 | +1.3 [−5.5, 8.2] | 0.53 | −0.55 [−1.40, 0.33] |
| HMC | 72 of 151 | WASO, min | +27.9 [−31.5, 87.3] | 0.70 | +35.9 [−31.2, 103.1] | 0.63 | +6.3 [2.0, 11.2] |
| | | Awakenings per night | +0.4 [−6.5, 7.4] | 0.68 | +1.1 [−4.1, 6.2] | 0.77 | −0.69 [−1.28, −0.14] |
| PhysioNet 2018 | 484 of 994 | WASO, min | +40.1 [−24.7, 105.0] | 0.57 | +31.2 [−27.6, 89.9] | 0.65 | −8.6 [−10.1, −7.0] |
| | | Awakenings per night | +3.9 [−7.6, 15.5] | 0.25 | +1.7 [−5.9, 9.2] | 0.44 | −1.94 [−2.28, −1.63] |
| BOAS PSG | 76 of 128 | WASO, min | +17.8 [−34.2, 69.7] | 0.63 | +22.1 [−25.6, 69.9] | 0.63 | +2.2 [−2.1, 6.1] |
| | | Awakenings per night | −1.7 [−8.2, 4.8] | 0.50 | −1.4 [−8.3, 5.6] | 0.47 | −0.03 [−0.58, 0.48] |
| CAP | 73 of 103 | WASO, min | +19.4 [−44.5, 83.3] | 0.74 | +30.2 [−45.1, 105.6] | 0.65 | +7.2 [3.1, 11.7] |
| | | Awakenings per night | +2.9 [−7.2, 13.1] | 0.08 | +3.2 [−4.8, 11.1] | 0.19 | −0.26 [−0.99, 0.44] |
| BOAS headband | 76 of 128 | WASO, min | +24.2 [−61.8, 110.3] | 0.37 | +31.8 [−54.3, 117.9] | 0.36 | +5.7 [−0.6, 12.5] |
| | | Awakenings per night | −3.1 [−9.3, 3.1] | 0.33 | −1.0 [−7.5, 5.5] | 0.50 | −1.09 [−1.70, −0.46] |
| Zmax headband | 69 of 96 | WASO, min | +39.7 [−135.6, 215.1] | 0.09 | +41.0 [−113.6, 195.5] | 0.14 | −1.8 [−15.0, 12.1] |
| | | Awakenings per night | −2.7 [−7.8, 2.4] | 0.25 | −0.7 [−6.4, 5.0] | 0.44 | −0.65 [−1.10, −0.14] |

Nights: nights whose first scored sleep onset followed the wake at lights-off and on which the device also marked onset, of all test nights. Device WASO: time in the awake state from the device's sleep onset to the last scored sleep epoch, so N1 and REM after an awake call count as wake. Bias: device minus scored value, with 95% limits of agreement (LoA). ICC: intraclass correlation, two-way random effects, absolute agreement, single measure. Difference in MAE: mean absolute error of WakeSentry minus that of GSSC, with 2,000 paired bootstrap resamples of persons; negative values favour WakeSentry.

**Table S3.** Awakening sensitivity by subgroup at the operating points fixed on the training cohorts.

| Subgroup | Persons | Scored awakenings | GSSC, % (FA/h) | WakeSentry, % (FA/h) | Difference, pp [95% CI] |
|---|---|---|---|---|---|
| **Age, y (Sleep-EDF, PhysioNet 2018, BOAS PSG, CAP)** | | | | | |
| <40 | 286 | 2,113 | 53.7 (0.37) | 58.7 (0.27) | +5.0 [2.5, 7.4] |
| 40–59 | 508 | 3,833 | 66.9 (0.63) | 65.2 (0.36) | −1.6 [−3.3, 0.1] |
| ≥60 | 477 | 4,205 | 60.0 (0.60) | 63.4 (0.39) | +3.4 [1.6, 5.3] |
| **Sex (Sleep-EDF, PhysioNet 2018, BOAS PSG, CAP)** | | | | | |
| Female | 469 | 3,784 | 62.1 (0.44) | 65.0 (0.31) | +2.9 [0.9, 4.9] |
| Male | 803 | 6,374 | 60.8 (0.63) | 62.0 (0.37) | +1.2 [−0.2, 2.5] |
| **Sex (BOAS and Zmax headbands)** | | | | | |
| Female | 124 | 1,021 | 36.4 (0.17) | 47.7 (0.24) | +11.3 [7.8, 14.8] |
| Male | 72 | 631 | 33.6 (0.17) | 52.5 (0.33) | +18.9 [13.6, 24.2] |
| **Apnoea–hypopnoea index, events/h (PhysioNet 2018)** | | | | | |
| <5 | 140 | 955 | 66.2 (0.69) | 65.0 (0.33) | −1.2 [−4.1, 1.5] |
| 5–15 | 287 | 2,076 | 69.9 (0.65) | 66.1 (0.33) | −3.8 [−5.9, −1.7] |
| 15–30 | 420 | 3,178 | 64.5 (0.77) | 62.9 (0.41) | −1.6 [−3.2, 0.1] |
| ≥30 | 145 | 1,076 | 60.5 (0.63) | 61.8 (0.43) | +1.3 [−1.6, 4.1] |
| **Diagnosis (CAP)** | | | | | |
| No disorder | 16 | 31 | 64.5 (0.24) | 80.6 (0.20) | +16.1 [4.5, 33.4] |
| Nocturnal frontal lobe epilepsy | 37 | 172 | 62.8 (0.21) | 70.3 (0.20) | +7.6 [3.2, 11.8] |
| REM sleep behaviour disorder | 21 | 94 | 50.0 (0.33) | 59.6 (0.37) | +9.6 [3.3, 16.5] |
| Periodic leg movements | 10 | 46 | 78.3 (0.65) | 73.9 (0.43) | −4.3 [−12.0, 4.0] |
| Insomnia | 9 | 33 | 63.6 (0.51) | 63.6 (0.51) | 0.0 [−20.0, 21.1] |
| Narcolepsy | 5 | 16 | 68.8 (0.40) | 68.8 (0.32) | 0.0 [−12.5, 20.0] |
| Sleep-disordered breathing | 4 | 9 | 22.2 (2.03) | 77.8 (1.20) | +55.6 — |
| Bruxism | 1 | 6 | 16.7 (0.00) | 50.0 (0.00) | +33.3 — |

Sensitivity at the operating points fixed on the training cohorts: detected over scored awakenings, pooled within the subgroup. FA/h: false awakenings per hour of stable sleep. Age was not available for HMC and for 4 other persons, sex for HMC and 3 CAP persons, and the apnoea–hypopnoea index for 2 persons. Apnoea–hypopnoea index: scored respiratory events per hour of sleep. CIs: 2,000 paired bootstrap resamples of persons; —, fewer than five persons. pp, percentage points.

**Table S4.** Awakening sensitivity by length of the wake bout and by the stage before waking.

| Awakenings | Measure | Sleep-EDF | HMC | PhysioNet 2018 | BOAS PSG | CAP | BOAS headband | Zmax headband |
|---|---|---|---|---|---|---|---|---|
| 1 epoch (30 s) | Scored awakenings | 543 | 555 | 3,397 | 734 | 56 | 734 | 222 |
| | GSSC → WakeSentry, % | 20.6 → 44.9 | 35.0 → 49.0 | 50.0 → 42.1 | 35.3 → 38.3 | 25.0 → 35.7 | 26.0 → 38.6 | 12.2 → 23.0 |
| | Difference [95% CI] | +24.3 [19.2, 29.3] | +14.1 [9.3, 19.2] | −7.9 [−9.9, −6.1] | +3.0 [−1.4, 7.2] | +10.7 [−1.8, 23.2] | +12.5 [8.2, 17.1] | +10.8 [5.9, 16.1] |
| 2–3 epochs | Scored awakenings | 338 | 307 | 1,651 | 244 | 157 | 244 | 159 |
| | GSSC → WakeSentry, % | 45.3 → 69.5 | 65.8 → 80.8 | 70.3 → 76.2 | 70.1 → 84.0 | 56.1 → 67.5 | 60.2 → 77.9 | 21.4 → 46.5 |
| | Difference [95% CI] | +24.3 [18.4, 30.7] | +15.0 [10.7, 19.5] | +5.9 [4.1, 7.8] | +13.9 [8.2, 20.4] | +11.5 [4.5, 18.4] | +17.6 [11.2, 24.3] | +25.2 [18.0, 33.6] |
| ≥4 epochs | Scored awakenings | 424 | 306 | 2,238 | 184 | 194 | 184 | 109 |
| | GSSC → WakeSentry, % | 80.7 → 81.1 | 83.0 → 86.3 | 85.9 → 88.1 | 87.0 → 91.3 | 74.2 → 78.4 | 77.7 → 85.9 | 38.5 → 56.9 |
| | Difference [95% CI] | +0.5 [−4.0, 4.8] | +3.3 [−0.6, 7.4] | +2.1 [1.1, 3.2] | +4.3 [0.0, 8.8] | +4.1 [−0.5, 8.5] | +8.2 [3.7, 13.7] | +18.3 [10.3, 26.4] |
| From N1 | Scored awakenings | 233 | 264 | 1,828 | 27 | 23 | 27 | 41 |
| | GSSC → WakeSentry, % | 50.2 → 56.7 | 53.8 → 65.9 | 54.0 → 54.0 | 55.6 → 51.9 | 56.5 → 52.2 | 48.1 → 59.3 | 31.7 → 48.8 |
| | Difference [95% CI] | +6.4 [−0.0, 12.9] | +12.1 [7.2, 17.2] | 0.0 [−1.8, 1.9] | −3.7 [−20.0, 13.0] | −4.3 [−28.6, 22.2] | +11.1 [−7.7, 29.0] | +17.1 [7.1, 30.0] |
| From N2 | Scored awakenings | 614 | 466 | 3,625 | 817 | 204 | 817 | 265 |
| | GSSC → WakeSentry, % | 55.4 → 72.0 | 67.8 → 71.5 | 75.6 → 70.7 | 59.7 → 62.2 | 64.7 → 72.5 | 49.8 → 59.6 | 22.6 → 38.9 |
| | Difference [95% CI] | +16.6 [12.8, 20.5] | +3.6 [−0.4, 8.2] | −4.9 [−6.5, −3.3] | +2.4 [−1.5, 6.8] | +7.8 [2.6, 13.6] | +9.8 [5.5, 14.4] | +16.2 [11.5, 21.6] |
| From N3 | Scored awakenings | 120 | 172 | 560 | 39 | 98 | 39 | 58 |
| | GSSC → WakeSentry, % | 29.2 → 56.7 | 55.8 → 74.4 | 64.5 → 68.9 | 30.8 → 69.2 | 58.2 → 74.5 | 23.1 → 56.4 | 27.6 → 60.3 |
| | Difference [95% CI] | +27.5 [20.9, 34.9] | +18.6 [10.3, 26.9] | +4.5 [1.1, 7.9] | +38.5 [22.2, 53.9] | +16.3 [10.0, 23.4] | +33.3 [18.2, 50.0] | +32.8 [19.4, 45.8] |
| From REM | Scored awakenings | 338 | 266 | 1,273 | 279 | 82 | 279 | 126 |
| | GSSC → WakeSentry, % | 34.0 → 53.6 | 36.1 → 56.0 | 54.6 → 56.8 | 26.9 → 37.6 | 53.7 → 54.9 | 18.6 → 38.0 | 11.1 → 23.0 |
| | Difference [95% CI] | +19.5 [13.4, 25.9] | +19.9 [15.1, 25.3] | +2.2 [−0.3, 4.6] | +10.8 [5.3, 16.4] | +1.2 [−7.5, 9.8] | +19.4 [13.2, 25.7] | +11.9 [5.2, 19.6] |

Sensitivity at the operating points fixed on the training cohorts: detected over scored awakenings. Length: number of 30-s wake epochs in the bout. Stage: the scored stage of the epoch before the bout. CIs: 2,000 paired bootstrap resamples of persons. Differences were computed before rounding.

**Table S5.** WakeSentry against GSSC with other dwell and hold times of the decision rule.

| Setting | Measure | Sleep-EDF | HMC | PhysioNet 2018 | BOAS PSG | CAP | BOAS headband | Zmax headband |
|---|---|---|---|---|---|---|---|---|
| Main analysis: 15 s, 9 s, 30 s | Sensitivity difference [95% CI] | +16.6 [13.3, 19.7] | +11.5 [8.5, 14.4] | −1.7 [−2.8, −0.6] | +5.5 [1.9, 9.3] | +7.9 [4.0, 11.6] | +12.9 [9.2, 17.0] | +17.1 [13.3, 21.1] |
| | FA/h, GSSC → WakeSentry | 0.16 → 0.40 | 0.32 → 0.29 | 0.70 → 0.37 | 0.21 → 0.15 | 0.36 → 0.31 | 0.18 → 0.23 | 0.16 → 0.33 |
| Asleep dwell 9 s | Sensitivity difference [95% CI] | +17.1 [13.8, 20.3] | +11.6 [8.6, 14.5] | −2.6 [−3.7, −1.5] | +5.8 [2.2, 9.4] | +7.6 [3.7, 11.4] | +13.1 [9.3, 17.2] | +17.6 [13.6, 21.6] |
| | FA/h, GSSC → WakeSentry | 0.16 → 0.41 | 0.32 → 0.29 | 0.72 → 0.36 | 0.21 → 0.15 | 0.37 → 0.31 | 0.17 → 0.23 | 0.16 → 0.33 |
| Asleep dwell 21 s | Sensitivity difference [95% CI] | +15.9 [12.6, 19.0] | +11.9 [8.9, 14.9] | −1.9 [−3.0, −0.8] | +5.2 [1.7, 8.9] | +8.6 [4.6, 12.4] | +13.0 [9.2, 17.2] | +17.3 [13.3, 21.5] |
| | FA/h, GSSC → WakeSentry | 0.16 → 0.40 | 0.31 → 0.29 | 0.70 → 0.37 | 0.21 → 0.15 | 0.35 → 0.32 | 0.18 → 0.23 | 0.16 → 0.33 |
| Awake dwell 6 s | Sensitivity difference [95% CI] | +5.7 [3.0, 8.4] | +10.0 [7.0, 13.0] | +3.0 [1.8, 4.1] | +5.4 [2.0, 8.9] | +6.9 [3.0, 10.9] | +11.3 [7.2, 15.6] | +14.9 [11.1, 19.0] |
| | FA/h, GSSC → WakeSentry | 0.20 → 0.22 | 0.38 → 0.35 | 0.58 → 0.41 | 0.26 → 0.18 | 0.38 → 0.36 | 0.23 → 0.27 | 0.22 → 0.40 |
| Awake dwell 12 s | Sensitivity difference [95% CI] | +8.7 [6.1, 11.3] | +4.3 [1.3, 7.1] | +6.3 [5.3, 7.4] | −4.5 [−7.2, −1.6] | +2.9 [−0.8, 6.7] | +2.8 [−1.0, 6.7] | +8.0 [4.5, 11.5] |
| | FA/h, GSSC → WakeSentry | 0.23 → 0.34 | 0.46 → 0.24 | 0.63 → 0.56 | 0.28 → 0.13 | 0.43 → 0.27 | 0.28 → 0.16 | 0.27 → 0.27 |
| Hold 15 s | Sensitivity difference [95% CI] | +16.8 [13.5, 20.0] | +11.5 [8.5, 14.4] | −1.7 [−2.8, −0.7] | +5.5 [1.9, 9.3] | +7.9 [4.0, 11.6] | +13.0 [9.3, 17.2] | +17.1 [13.3, 21.1] |
| | FA/h, GSSC → WakeSentry | 0.16 → 0.40 | 0.32 → 0.29 | 0.72 → 0.38 | 0.21 → 0.15 | 0.37 → 0.31 | 0.18 → 0.23 | 0.16 → 0.33 |
| Hold 60 s | Sensitivity difference [95% CI] | +16.5 [13.1, 19.6] | +11.4 [8.4, 14.4] | −1.3 [−2.4, −0.2] | +5.3 [1.7, 9.0] | +7.9 [4.0, 11.7] | +12.8 [9.0, 17.0] | +16.9 [13.1, 20.8] |
| | FA/h, GSSC → WakeSentry | 0.16 → 0.40 | 0.31 → 0.29 | 0.67 → 0.37 | 0.20 → 0.15 | 0.34 → 0.30 | 0.17 → 0.23 | 0.16 → 0.33 |

Each setting changes one of the dwell time for ‘asleep’, the dwell time for ‘awake’ and the hold time; operating points of both methods were chosen again on the training cohorts. Sensitivity difference: WakeSentry minus GSSC, percentage points, at the operating points. FA/h: false awakenings per hour of stable sleep. CIs: 2,000 paired bootstrap resamples of persons.
